# SGLT2-Inhibition reverts urinary peptide changes associated with severe COVID-19: an *in-silico* proof-of-principle of proteomics-based drug repurposing

**DOI:** 10.1101/2021.07.21.21260351

**Authors:** Agnieszka Latosinska, Justyna Siwy, David Z. Cherney, Bruce A. Perkins, Harald Mischak, Joachim Beige

## Abstract

Severe COVID-19 is reflected by significant changes in multiple urine peptides. Based on this observation, a clinical test based on urinary peptides predicting COVID-19 severity, CoV50, was developed and registered as IVD in Germany. We have hypothesized that molecular changes displayed by CoV50, to a large degree likely reflective of endothelial damage, can be significantly reversed by specific drugs. To test this hypothesis, we have collected urinary peptide data from patients without COVID-19 prior and after drug treatment. The drugs chosen were selected based on availability of sufficient number of participants in the dataset (n>20) and potential value of drug therapies in the treatment of COVID-19 based on reports in the literature. In these participants without COVID-19, while spironolactone did not demonstrate a significant impact on CoV50 scoring, empagliflozin treatment resulted in a significant change in CoV50 scoring, indicative of a potential therapeutic benefit. The results serve as a proof-of-principle for a drug repurposing approach based on human urinary peptide signatures and support the initiation of a randomised control trial testing a potential positive effect of empagliflozin in the treatment of severe COVID-19, possibly via endothelial protective mechanisms.

**Significance of the study:** COVID-19 pandemic has imposed a heavy burden on society, health care and economics. Although multiple drugs have been tested in the context of COVID-19, effective treatments for patients experiencing severe disease are still missing, with some drugs demonstrating benefit only at earlier disease stage. Computational drug repurposing emerged as a promising approach to boost drug development, allowing to predict drug efficacy based on the molecular signature of drug impact, mainly using transcriptomics data from cell lines.

Recently we demonstrated that urinary proteomics profiles significantly differ between patients with severe COVID-19 course and those with mild/ moderate disease. This resulted in the development of a molecular signature associated with COVID-19 severity (CoV50), allowing to predict COVID-19 course, and enabling guiding intervention.

Here we report on the first study demonstrating the application of clinical proteomics data (from clinical trial participants) in a drug repurposing approach. We used the CoV50 signature to examine if the molecular changes associated with COVID-19 severity in patients without COVID-19 might be altered by existing drugs. In a study population without COVID-19, empagliflozin demonstrated a partial, yet significant reversion of the CoV50 signature, indicating a potential benefit in the context of severe COVID-19.

## Main text

The current COVID-19 pandemic has generated multiple challenges for clinicians and patients. One major issue is that severity of disease course cannot be predicted with good certainty at an early point in time. While most patients experience a moderate course of disease, comparable with common cold, a fraction of the infected population develops severe symptoms that lead to requirement of oxygenation, organ failure and death in a significant number of subjects in this group ^[1]^. Risk factors for severe COVID-19 are, among others, age, presence of other diseases as diabetes, obesity and higher stages of chronic kidney disease ^[2]^. While these risk factors allow prediction of disease course with some confidence, they are not linked to molecular pathophysiology. In addition, these risk factors are generally not modifiable, limiting their value in guiding intervention. Therapeutic interventions in severe COVID-19, generally separated in (early) anti-replicative therapies and (later) therapies tackling dysregulated immune response have not yielded major outcome improvements in a number of controlled trials ^[3]^. Beside the primary virus entry mechanism and virus replication, meaningful pathways serving as targets for interventions tackling the indirect (later) diseases sequelae are lacking.

In a recent study we have investigated urine peptides for their association with severe COVID-19 ^[4]^. Positive preliminary data resulted in the initiation of a prospective multicentre clinical study. Patients with severe COVID-19 course and patients with mild/ moderate COVID-19 have a substantially different urinary peptide pattern (UPP) based on which a classifier termed CoV50 was developed ^[5]^, representing a molecular signature associated with disease severity ^[5]^. Due to the urgent need presented by the pandemic, an interim assessment of the study was initiated and recently published, demonstrating that CoV50 enables prediction of COVID-19 severity, and consequently early intervention ^[5]^. Based on the data, the CoV50 test was approved on the basis of the Directive 98/79/EC on In-Vitro Diagnostic Devices by the Federal Institute for Drugs and Medical Devices (BfArM), the responsible German authority (registration no: DE/CA09/0829/IVD/007) and is being used to predict COVID-19 course, aiming at improving outcome with early therapeutic intervention.

A detailed investigation of the urine peptides significantly affected in patients that subsequently develop severe COVID-19 indicated highly significant similarity to peptides altered in chronic and acute kidney disease, while no similarity to peptides altered in heart failure was detectable ^[6]^. The results of these investigations also indicated that severe COVID-19 (defined throughout this manuscript as World Health Organization (WHO) grade 6 or higher) may be associated with endothelial damage, which is in agreement with the current literature ^[7]^. One of the consequences of such a hypothesis would be that therapy directed against SARS-CoV-2 is not expected to deliver benefit if initiated at a later stage of the disease, when the endothelial layer is already damaged due to the virus infection. In fact, it has been shown that antiviral therapies display benefit only if initiated in the very early phases of SARS-CoV-2 infection ^[8.9]^.

Based on these data, intervention targeting the damages that result from the viral infection, especially in the endothelium (being consequence of and detectable in severe COVID-19), and not the SARS-CoV-2 virus *per se*, may be beneficial in the treatment/prevention of severe COVID-19.

Drug repurposing has gained significant interest especially from pharmaceutical industry, mainly due to the opportunity to decrease overall cost of developing new drugs and timelines ^[10]^. One computational (*in silico)* approach that has been used by multiple groups ^[11]^ including us ^[12.13]^, is prediction of drug efficacy based on the molecular signature of drug impact ^[14]^. One of the well-established solutions in this respect is the Connectivity Map (CMap) analysis ^[15]^ in which molecular disease signatures are matched against the transcriptomic-based molecular signatures induced by drug treatment *in vitro*.

Given the knowledge on the molecular signature associated with COVID-19 severity, as well as access to urinary peptide data from interventional studies involving patients without COVID-19, all based on the same technology and highly comparable ^[16]^, in this study we aimed towards investigating if any of the drugs assessed in the different studies does result in a significant change of the severe COVID-19 urinary peptide signature (CoV50), in analogy to the CMap approach.

The outline of this study is graphically depicted in **Figure 1**. When aiming at identifying a potential drug benefit in the context of COVID-19, ideally patients with COVID-19 receiving drug or placebo should be investigated. Unfortunately, data on the drugs in COVID-19 are generally not available. To predict if drugs may be beneficial in this context, the impact of drugs on the CoV50 signature can be investigated based on data from subjects without COVID-19 before and after drug treatment. Samples from these patients are expected to score negative for CoV50 signature, since they did not have COVID-19. A change in the positive direction would be considered a signal for a potential negative effect of the drug, a change in the negative direction (reverting the CoV50 signature) would be considered a signal for a potential benefit. Similar approaches have been used mostly based on transcriptomics data and cell lines, with one of the most prominent approaches being the CMap ^[17]^. The approach chosen here is the first report on such efforts based on real patient data, using urinary peptides, supporting the use of human-derived data for drug repurposing.

**Figure 1:**
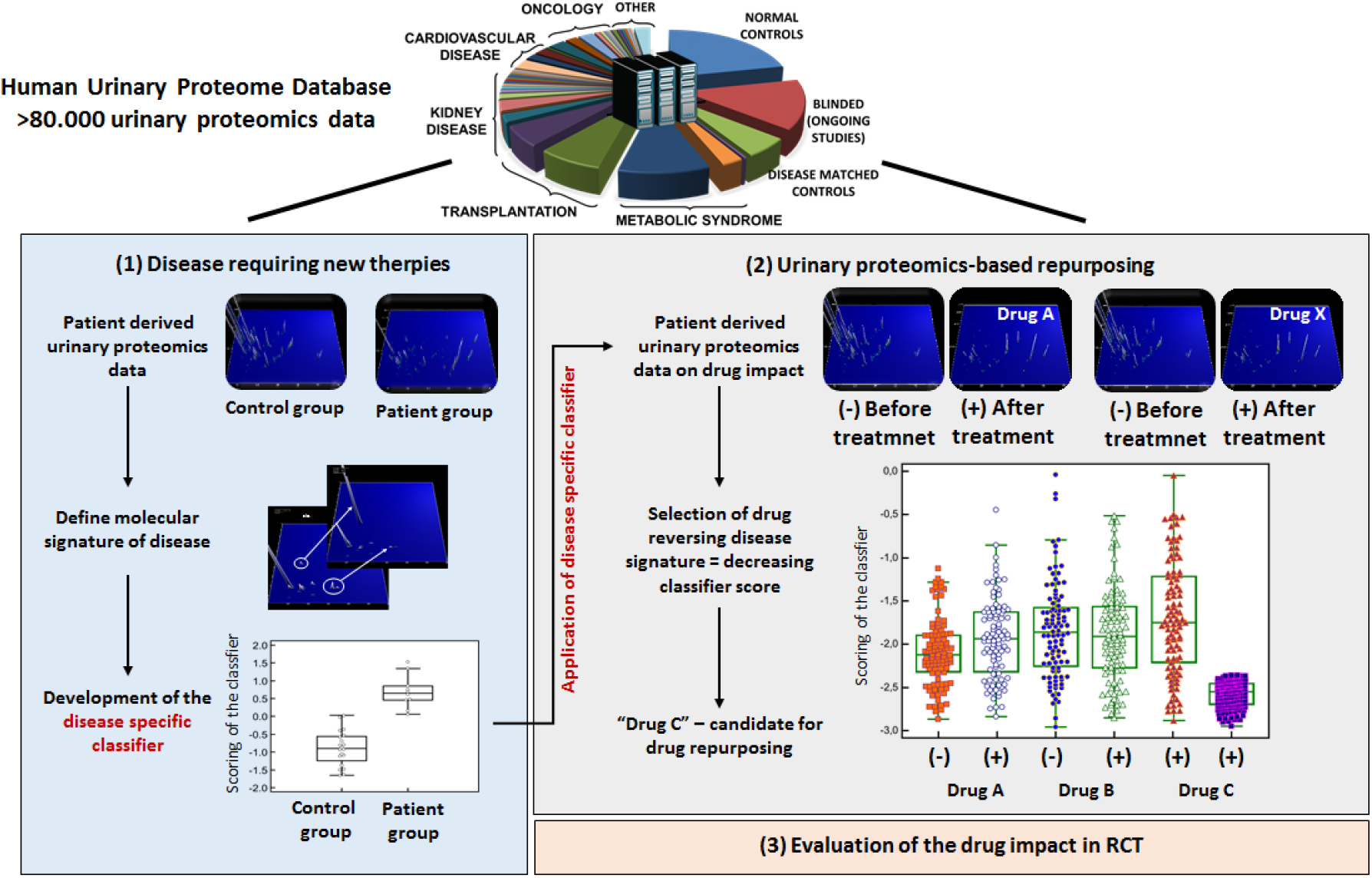
Schematic depiction of the study design. The underlying hypothesis of the study was that drugs that are able to revert a disease signature, here a signature based on urine peptides, may be beneficial in treating the respective disease. As such, we tested if a urinary peptide-based classifier, CoV50, that is associated and predicts critical course of COVID-19 (left panel), is affected by drug treatment. For this purpose, data from COVID-19 free patients before and after treatment with specific drugs from the human urinary database were extracted and the impact of the drug treatment on the CoV50 scoring was investigated. **Abbreviations**: RCT – randomised controlled trial.

Power calculations indicated that in order to detect a 50% change in the CoV50 scoring with a *p*-value <0.05 and 80% power, datasets from a minimum of 17 subjects before and after treatment are required. We therefore only included studies involving at least 20 subjects. Investigation of the data in Urine Proteome Database ^[16]^ resulted in the identification and extraction of datasets from patients without COVID-19 undergoing treatment with the aldosterone antagonist spironolactone from the HOMAGE trial ^[18]^, and the sodium-glucose transport protein 2 (SGLT2)-inhibitor empagliflozin ^[19.20]^. As a part of the HOMAGE trial ^[21]^, the placebo group was also investigated. The details on the studies and the patient data sets included are listed in **Table 1**.

**Table 1:**
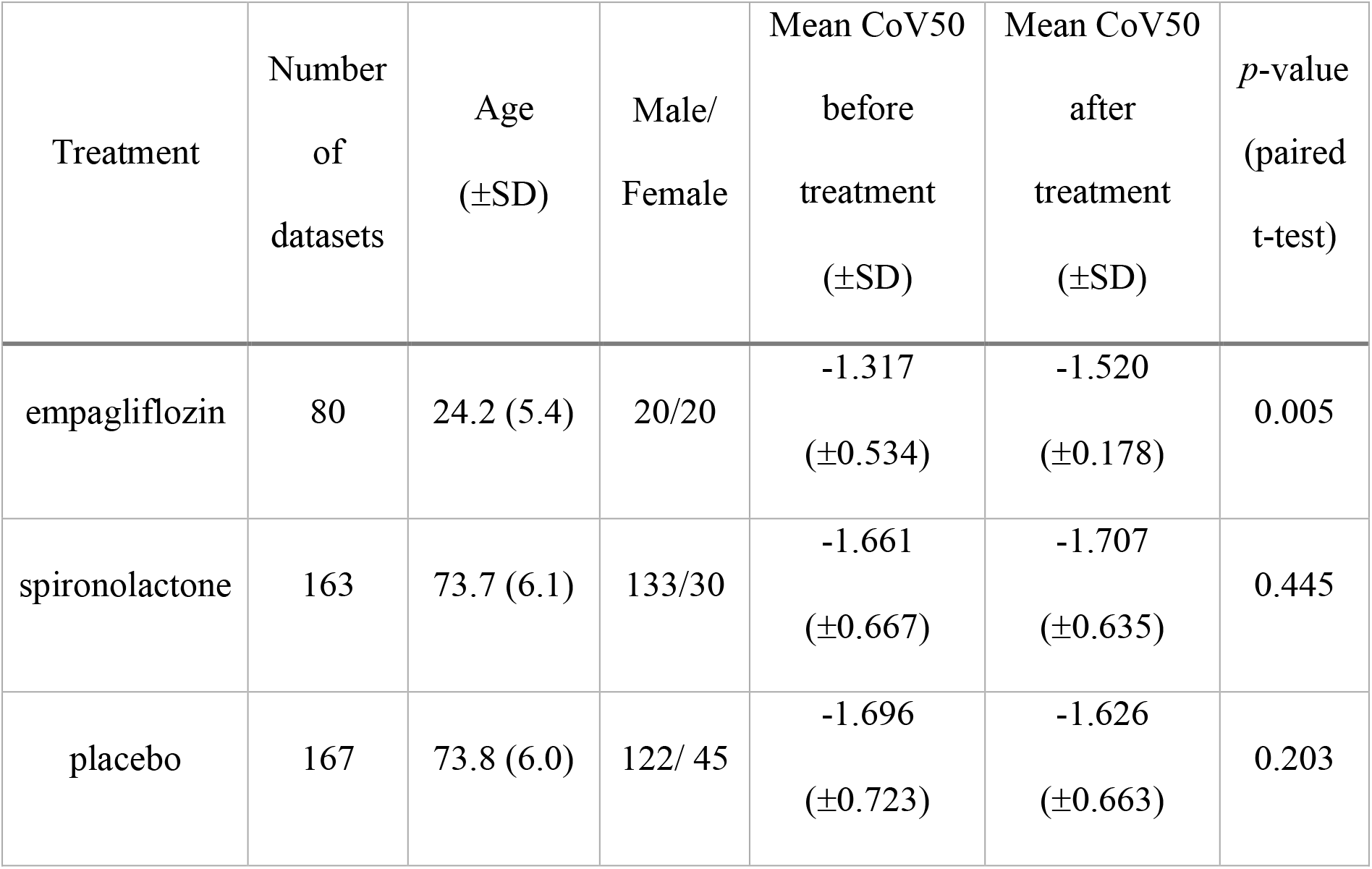
Characteristics of the subjects and datasets investigated in this study.

To investigate if an impact of the treatment with the respective drug on COVID-19 associated UPP-changes could be expected, the CoV50 classifier was applied on samples from patients without COVID-19 collected at baseline (before treatment) and during follow-up (after treatment). The distribution of the CoV50 scoring in the baseline and follow-up samples is shown in **Figure 2**. While this was a study in a population without COVID-19, participants had detectable levels of urinary peptides included in CoV50 signature. As expected in a study population without COVID-19, the study subjects generally scored negative for severe COVID-19 and there was no significant difference in the CoV50 scoring in the placebo group before and after treatment. No significant difference was also detectable as a result of treatment with spironolactone. In contrast, empagliflozin treatment significantly reduced the level of the CoV50 score in a dataset of 160 samples ^[20]^ (80 before and 80 after treatment with empagliflozin), with a significant decrease (*p*=0.04) from an average score of -1.76 (before treatment) to an average of -1.89 (after treatment). The observation that the molecular profile induced by severe COVID-19 opposes the changes induced by empagliflozin, even in patients without COVID-19, indicate that exposure to empagliflozin may revert, at least in part, the molecular changes in severe COVID-19.

**Figure 2:**
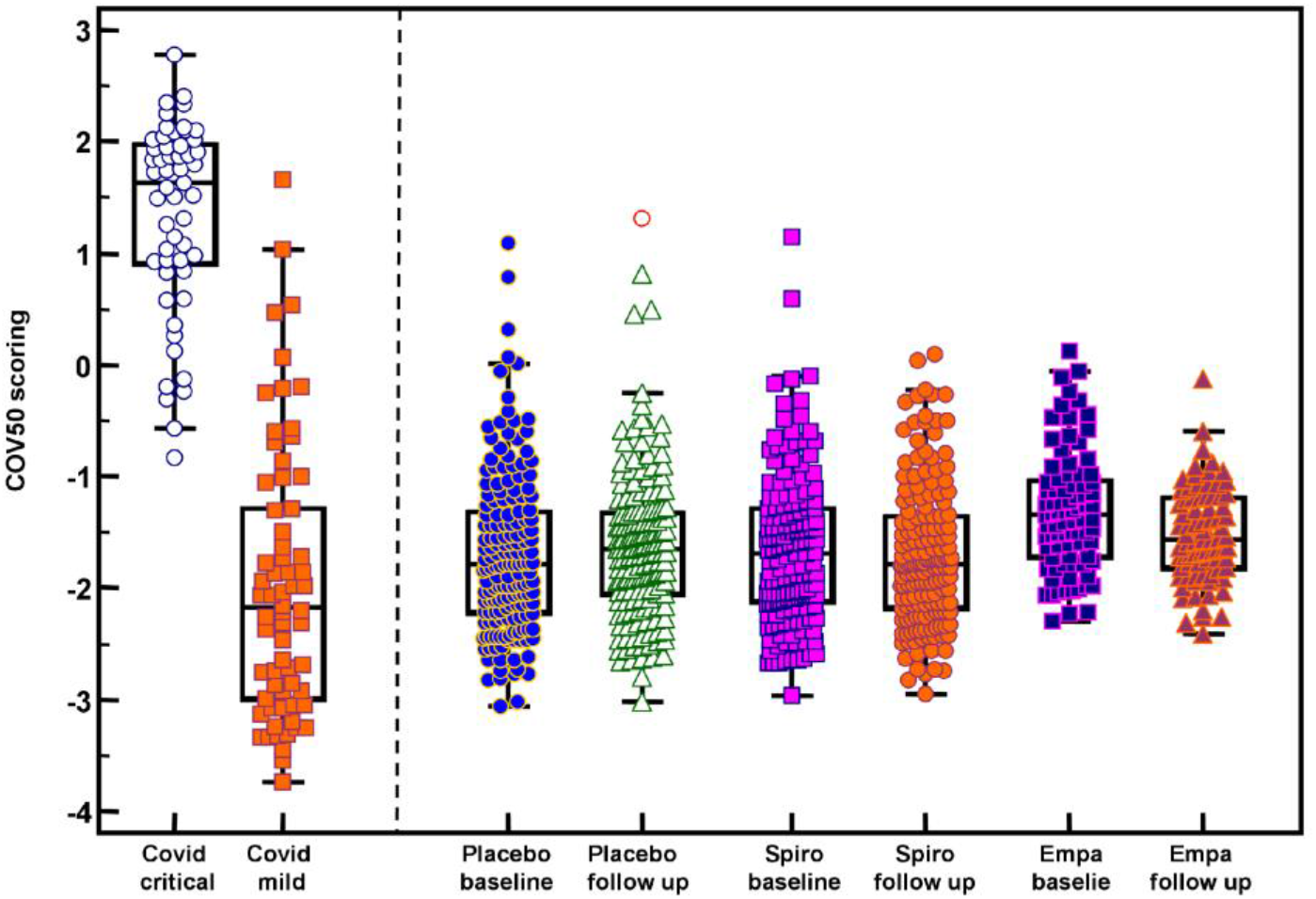
CoV50 scoring of the subjects investigated. For comparison, the distribution of CoV50 scoring of patients with mild (WHO grade 1-3) or critical (WHO grade 6-8) disease is presented on the left. **Abbreviations**: Empa – empagliflozin, and Spiro – spironolactone.

A significant difference in the CoV50 scoring between the spironolactone and the empagliflozin cohorts was observed. This may be due to the significant difference in age (see **Table 1**), however, we have not detected an association of CoV50 scoring with age. A more likely explanation could be the different trial locations: the HOMAGE trial was conducted in Europe, while the empagliflozin cohort is from Canada.

Therapeutics applied to COVID-19 in controlled trials comprise attempts of biological therapies (monoclonal anti-virus directed ^[22]^ and anti-C5a antibodies ^[23]^ and polyclonal convalescent plasma ^[24]^), inflammation inhibitors ^[25]^, small-molecule anti-virus therapies (remdesivir, lopinavir and ribavirin, hydroxychloroquine ^[26]^) and steroids ^[27]^ in high and medium dosage schemes. Several authors suggested spironolactone may be of potential positive impact in the context of COVID-19 ^[28]^. However, these suggestions were generally based on theoretical considerations, and we could not identify a clinical trial testing a potential benefit of spironolactone. In the BISCUIT trial a limited number of patients treated with bromohexine and spironolactone was compared to placebo, and a significant reduction on the duration of COVID-19 was reported ^[29]^. However, with 33 patients the study is very small, the fractional impact of any one of the two drugs apparently has not been assessed, consequently the impact of spironolactone treatment on COVID-19 cannot be evaluated with sufficient certainty. Our data presented here do not indicate a potential benefit of spironolactone.

We could not find published data from a study assessing the impact of SGLT2 inhibition on COVID-19 course. However, such data should soon become available from the DARE-19 study ^[30]^. Currently the most informative publication in this context may be the data published by Israelsen and colleagues who reported similar impact of SGLT2 inhibitors and glucagon-like peptide-1 agonists ^[31]^. Unfortunately, however, the study did not compare to patients taking neither drugs.

Our main finding, that SGLT2 inhibitors may be beneficial in the context of severe COVID-19, are further supported by reports in animal studies on protective effects of SGLT2 inhibition in the context of endothelial dysfunction ^[32]^. Since a major component of severe COVID-19 appears to be endothelial dysfunction, drugs that demonstrate protective effects in this context may display benefits in preventing/reducing severe COVID-19.

Our study has shortcomings. First, it is a retrospective analysis of previously collected data. Second, the impact of the different drugs on the CoV50 classifier cannot be interpreted as sure predictors of efficacy in COVID-19. This analysis was conducted in COVID-19 free patients and the CoV50 scores, as expected, did not indicate severe disease. Nevertheless, the observed significant decrease of the CoV50 score following treatment, suggests that SGLT2 inhibitors may confer some protection from severe COVID-19 course. A significant impact of SGLT2 inhibition on the course of COVID-19 would need to be demonstrated in a properly powered prospective clinical trial. However, the manuscript presented here is based on a substantial number of observations, and can certainly be interpreted as a significant supportive dataset to initiate testing the potential benefits of SGLT2 inhibition in COVID-19.

In conclusion, the study and data presented here are a first indication that clinical proteomics data from clinical trial participants can be exploited in a drug repurposing approach. Several studies reported on the repurposing of established drugs like aspirin ^[33]^ and statins ^[34]^ to abolish COVID-19 clinical thrombo-embolic endpoints, however, mostly in observational rather than randomised control trials. While proteomic data are usually not available from these observational studies, it should be feasible to identify non-COVID investigations with e.g. aspirin and statins, and *in-silico* model the consequences of such treatments, based on urinary proteomics/peptidomics, as demonstrated in this study. A substantial benefit of the approach shown here may be that it is based on patient data, not on cell lines or animal models, consequently may better depict pathophysiology in patients, which are by far more complex than single cells.

## Experimental Procedures

For this study urinary proteome data obtained by capillary electrophoresis coupled to mass spectrometry (CE-MS) and stored in the Human Urine Proteome Database ^[16]^ were assessed. This database currently contains urinary proteome/ peptidome information on >80.000 samples, assessing a dataspace comprised total of >100.000 peptides and low molecular weight proteins ^[16]^. Information on the peptide sequencing from liquid chromatography coupled to tandem mass spectrometry (LC-MS/MS) and CE-MS/MS analysis, and anonymized clinical information of participants enrolled in the studies are also included. The CE-MS technology applied has been described in detail including reproducibility, repeatability, procedures for sample preparation, data evaluation and normalization ^[35]^. More detailed information on the peptide sequencing was described elsewhere ^[36]^.

All underlying studies were conducted to conform to regulations on the protection of individuals participating in medical research and in accordance with the principles of the Declaration of Helsinki (2013) and received ethical approval by the responsible institutional review boards. Written informed consent was obtained from all participants at the time of sampling. All data sets received were anonymized. Specifically, for all samples from COVID-19 patients the Ethics Committee of the German-Saxonian Board of Physicians, Dresden, Germany (number EK-BR-88/20.1) and the Institutional Review Boards of the recruiting sites provided ethical clearance ^[5]^. The protocol was registered with the German Register for Clinical Studies (www.drks.de), number DRKS00022495, which is interconnected with the WHO International Clinical Trial Registry Platform (www.who.int/clinical-trials-registry-platform). For the samples from patients undergoing empagliflozin treatment, all patients consented to the study, which was approved by the University of Toronto University Health Network with approval #11-0213 ^[20]^. All samples from subjects undergoing spironolactone treatment and placebo controls were from the HOMAGE trial, a prospective, randomized, open-label, blinded-endpoint (PROBE), multicentre design, in which people at increased risk of developing HF were randomly assigned to receive either spironolactone or standard of care (“control”) - not receiving spironolactone or other mineralocorticoid receptor antagonists (ClinicalTrials.gov Identifier: NCT02556450). The study was approved by all relevant ethics committees and regulatory bodies ^[37]^.

Distribution of the CoV50 scoring was evaluated using MedCalc (version 12.1.0.0, MedCalc Software, Mariakerke, Belgium; https://www.medcalc.org/). For analysis of differences in the scoring between baseline and drug treatment, paired T testing was applied, statistical significance was assumed at *p* < 0.05.

## Data Availability

The data that support the findings of this study are available from the corresponding author upon reasonable request.

## Abbreviations

CMap: Connectivity Map
SGLT2: sodium-glucose transport protein 2
UPP: urinary peptide pattern
WHO: World Health Organization.

